# Characterization of the common genetic variation in the Spanish population of Navarre

**DOI:** 10.1101/2024.02.11.24302643

**Authors:** Alberto Maillo, Estefania Huergo, María Apellániz-Ruiz, Edurne UrruDa, María Miranda, Josefa Salgado, Sara Pasalodos-Sánchez, Luna Delgado-Mora, Óscar Teijido, Ibai Goicoechea, Rosario Carmona, Javier Perez-Florido, Virginia Aquino, Daniel Lopez-Lopez, María Peña-Chilet, Sergi Beltrán, Joaquín Dopazo, Iñigo Lasa, Juan José Beloqui, NAGEN-scheme, Ángel Alonso, David Gomez-Cabrero

## Abstract

**Purpose:** Large-scale genomic studies have significantly increased our knowledge of genetic variability across populations. Regional genetic profiling is essential for distinguishing common benign variants from disease-causing ones. To this end, we conducted a comprehensive characterization of exonic variants in the population of Navarre (Spain).

**Methods:** Genome sequencing data from 358 unrelated individuals of Spanish origin from the Navarrese population were used.

**Results:** Our analysis revealed 61,410 biallelic exonic single nucleotide variants (SNV) within the Navarrese cohort, with 35% classified as common (MAF > 1%). By comparing allele frequency data from 1000 Genome Project (excluding the Iberian cohort of Spain, IBS), Genome Aggregation Database, and a Spanish cohort (including IBS individuals and data from Medical Genome Project), we identified 1,069 SNVs common in Navarre but rare (MAF ≤ 1%) in all other populations. We further corroborated this observation with a second regional cohort of 239 unrelated exomes, which confirmed 676 of the 1,069 SNVs as common in Navarre.

**Conclusion:** This study highlights the importance of population-specific characterization of genetic variation to improve allele frequency filtering in sequencing data analysis to identify disease-causing variants.

## INTRODUCTION

In recent years, the use of NGS in patient healthcare has increased due to technological advances, cost reduction, and enhanced efficiency.^1^ The advancement of NGS spans a spectrum of applications, encompassing whole exome/genome sequencing (WES/WGS). These technologies revealed a wealth of genetic variants, necessitating the implementation of filters to narrow down the list of candidate variants. In this regard, the availability of population-specific catalogues of common variants enables the identification of rare variants ^2^, such as the international initiatives 1000 Genome Project (1KGP)^3^ and Genome Aggregation Database (gnomAD) ^4^. Moreover, various countries like the UK,^5^ USA,^6^ and Japan^7^ have established their databases. In Spain, for instance, the Medical Genome Project (MGP) compiles data from unrelated healthy individuals.^8,9^

In Navarre, a 650,000 population region of north-eastern Spain, the local Government supported the “*NAGEN scheme*” to integrate genomic data analysis into the regional public healthcare system. Nowadays, NAGEN has generated numerous WES/WGS and associated phenoclinical profiles in seven projects, including *NAGEN1000*, focused on rare diseases, and *pharmaNAGEN* on pharmacogenomics in patients with inflammatory bowel diseases.^10^ The NAGEN strategy’s success hinges on identifying population-specific common variants to establish a comprehensive Navarrese population frequency catalogue.

In this study (Fig. 1), we aimed to identify and characterize common exonic variants specific to the Navarrese population. Firstly, we identified common single nucleotide variants (SNVs) in Navarre and rare in other populations. Secondly, we validated the allele frequency of these variants in another Navarrese cohort with exome data. Finally, we annotated the resulting variants using genomic databases, and their clinical and pharmacological effects and pathogenicity were assessed. Additionally, we conducted functional enrichment analyses to provide further insights. The results will significantly contribute to advancing personalized medicine in Navarre.

**Figure 1:**
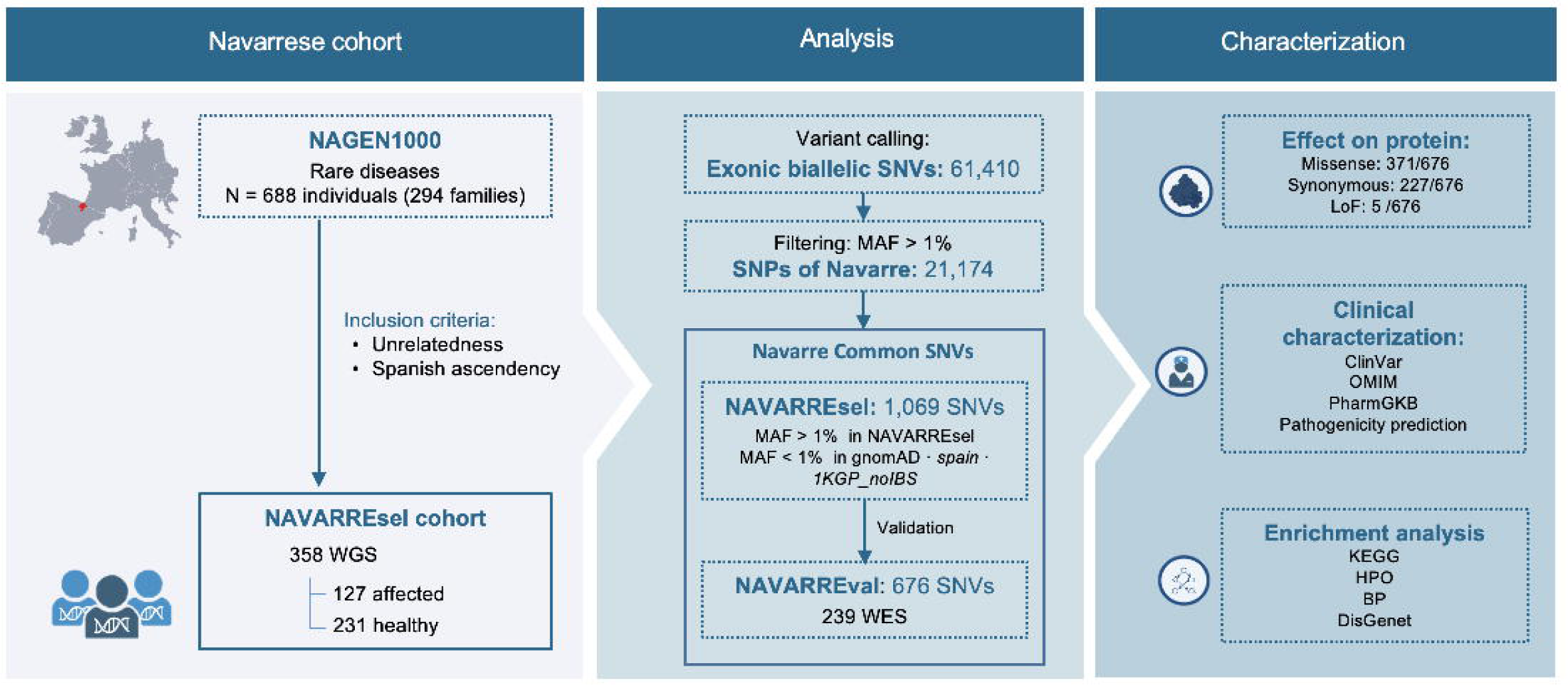
Workflow of this study. Abbreviations: *MGP*, Medical Genome Project; *1KGP*, 1000 Genomes Project; *1KGP_noIBS*, 1000 Genomes Project without Iberian population; *gnomAD,* Genome Aggregation Database; *MAF*, minor allele frequency; *SNV*, single nucleotide variant; *SNP*, single nucleotide polymorphism; *WGS*, whole genome sequencing; *WES*, whole exome sequencing; *LoF*, Loss-of-function; HPO, Human Phenotype Ontology; BP, biological process.

## MATERIAL AND METHODS

Please refer to the Supplementary information for detailed material and methods.

### Referenced population projects

Population frequencies were obtained from 1) gnomAD: genomes v2.1.1; 2) *1KGP_noIBS*: mean 1KGP (phase3) population’s frequencies, excluding the IBS cohort; and 3) *spain*: combining the MGP and IBS frequencies.

### Variant annotation

Variants were annotated using ANNOVAR.^11^ Clinical and pharmacological relevance was assessed using ClinVar,^12^ Online Mendelian Inheritance in Man (OMIM)^13^ and PharmGKB.^14^ Variant classification was conducted following the American College of Medical Genetics (ACMG) guidelines.^15^

## RESULTS

### Navarrese discovery cohort

The *NAGEN1000* WGS Navarrese project comprised 688 individuals from 294 families (mainly trios) with a rare disease. The WGS was conducted with a mean coverage of 30X, providing comprehensive genomic data across the entire genome. For our study, we kept a subset of this cohort satisfying two criteria: unrelatedness and Spanish ancestry. This result yielded 358 individuals, referred to as NAVARREsel.

Then, biallelic SNVs on chromosomes 1 to 22, from the exonic region covered by the Nextera Exome Enrichment kit, were extracted. Subsequently, variants with read depth < 10, genotype quality < 50, or missing genotype in at least one sample were filtered out. Additionally, sites significantly deviated from Hardy-Weinberg equilibrium (HWE, p-value < 10^-5^) were removed.^16^ Finally, 61,410 SNVs remained, of which 21,174 were identified as common variants (MAF > 1%). We observed that including additional individuals did not reveal new common variants, and 21,174 were achieved when considering over 100 individuals (Fig. S1a).

### Genetic variation between Navarre, Spanish, and global populations

We performed a principal component analysis (PCA) on the shared variants between NAVARREsel, 1KGP, and MGP to depict its relationship. We observed a clear distinction between Navarre and Asian/African populations, reflecting established genetic differences (Fig. 2a). Conversely, an overlap is observed between Navarre and European populations, emphasizing their genetic affinity. Thus, focusing on European populations (Fig. 2b), we observed that Navarrese individuals are close to the Spanish populations (IBS and MGP) and exhibit proximity to Italian individuals.

**Figure 2:**
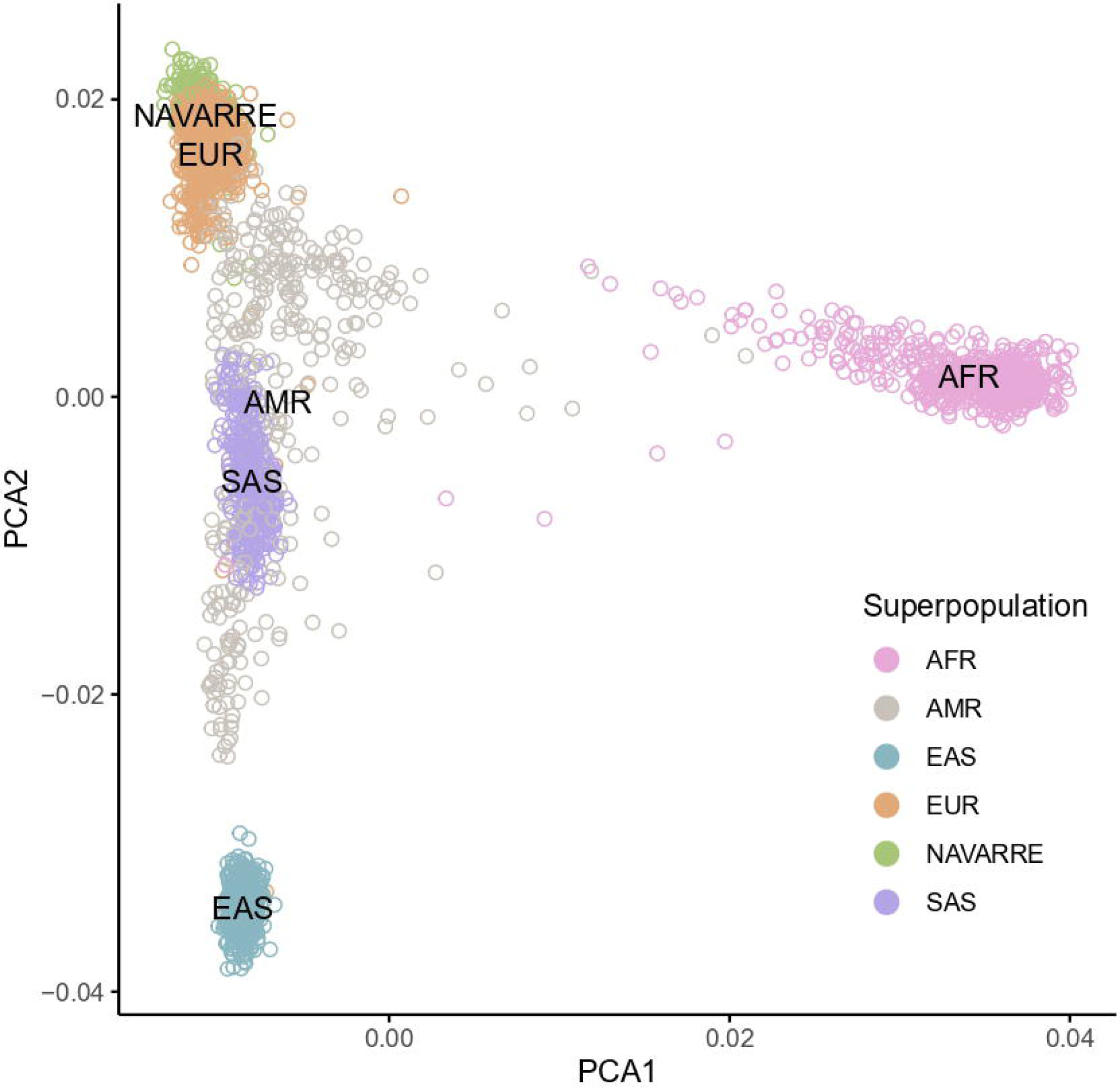
**a)** Principal component analysis of overlapped variants between NAVARREsel, MGP, and 1KGP (including all populations), and coloured by superpopulations. **b)** Principal component analysis of overlapped variants between NAVARREsel, MGP, and 1KGP (including exclusively European populations). **c)** Genetic admixture analysis of 1,128 individuals from 7 European populations for the optimal *K* value = 3. Abbreviations: PCA, principal component analysis; AFR, African populations; AMR, American populations; EAS, east-Asian populations; SAS, south-Asian populations; EUR, European populations; IBS, Iberian populations in Spain; MGP, Medical Genome Project; TSI, Toscani in Italy; CEU, Utah residents with Northern and Western European ancestry; GBR, British in England and Scotland; FIN, Finnish in Finland.

This observation is supported when estimating the ancestries of the European populations using ADMIXTURE.^17^ The average number of ancestries in each population was calculated with the optimal component K=3 (Fig. 2c). The Navarrese population showed the highest ancestral proportion on component 1 at 61%, which started decreasing in the IBS and MGP populations to 30% and 20% respectively, and was nearly absent (0.1%) in Finland (FIN). In contrast, component 2 was predominant in the Finnish population (99%), while being the lowest in the Navarrese cohort at 7%.

To further analyse the genetic differentiation, we calculated the mean pairwise F_ST_ values. The lower F_ST_ value indicates greater similarity between populations. This occurred when comparing Navarre with the Spanish (F_ST(Navarre-IBS)_ = 0.0001 and F_ST(Navarre-MGP)_ = 0.0007) and Italian (F_ST(Navarre-TSI)_ = 0.0014) populations. In contrast, the highest differentiation was observed against East-Asian and African populations (F_ST(Navarre-EAS)_ = 0.0328, F_ST(Navarre-AFR)_ = 0.0434, Table S1).

These findings, aligning with biological expectations, underscore the regional and continental genetic affinities, providing insights into historical populations and evolutionary dynamics.

### Exclusive common variants in Navarre

To identify exclusive Navarrese common variants, we examined allele frequency among Navarre population and the three referenced populations: *1KGP_noIBS*, gnomAD, and *spain*. A comparison of the MAF revealed that most variants (17,532 SNVs) were classified as common (MAF > 1%) across the four populations. However, 835 variants exhibited higher prevalence solely in Spanish cohorts (Navarre and *spain*). Specifically, 1,069 SNVs were identified as common in Navarre, and rare (MAF ≤ 1%) in the rest (Fig. 3a).

**Figure 3:**
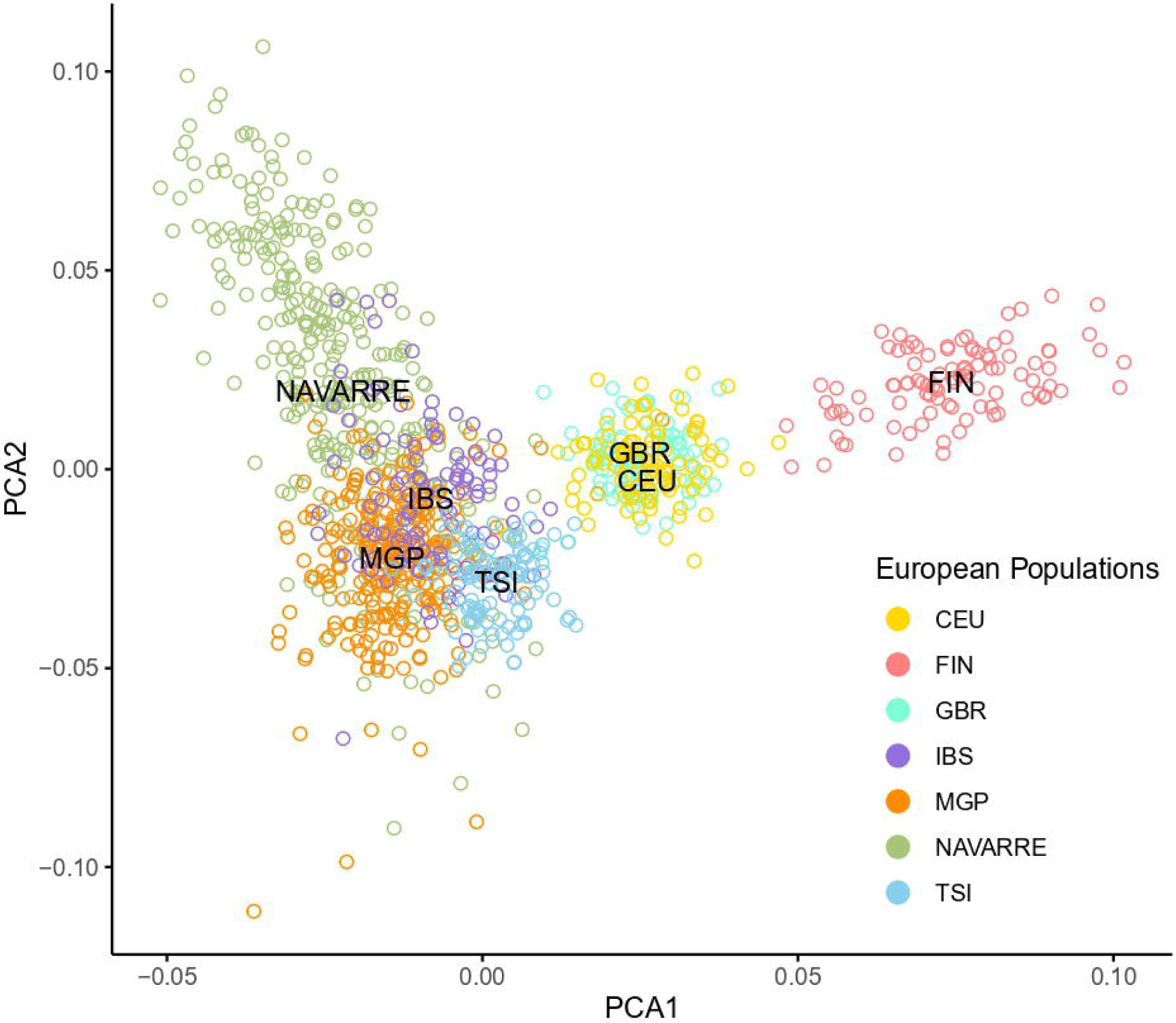
**a)** Upset plot of common variants (MAF > 1%) of each population: NAVARREsel, *spain*, *1KGP_noIBS*, and gnomAD **b)** Resulting percentage of variants per patient (n=127) after removing common variants from Navarre, *spain*, gnomAD, or *1KGP_noIBS* populations. The box plots represent the median, upper, and lower quartiles by the centre line and box bounds, respectively. Whiskers display the largest and smallest values within 1.5 times the interquartile range from the quartiles. Abbreviations: *1KGP_noIBS*, 1000 Genomes Project without Iberian population; gnomAD, Genome Aggregation Database; *spain*, integration of IBS and MGP populations.

To validate these 1,069 variants, we used the NAVARREval cohort, a subset of 239 WES unrelated individuals of Spanish descent from the current Navarrese population and diagnosed with Crohn’s disease (159/239) or ulcerative colitis (86/239) (*pharmaNAGEN* project). Before validation, we assessed the association of these SNVs with these conditions by cross-referencing them with reported variants in the Inflammatory Bowel Disease database, which catalogues variants highly linked to the mentioned diseases.^18^ The absence of the 1,069 SNVs in this database ensured an unbiased and robust validation process.

Among the 1,069 variants initially identified, 998 were detected in NAVARREval with a call rate greater than 80% and demonstrated conformity to HWE. Notably, 676/998 of these SNVs (68%; p-value = 2.2e^-16^) were consistently classified as common in NAVARREval, confirming their prevalence within the Navarrese population (variants’ information in Table S2). The validation cohort was sufficient to validate the Navarrese common variants, reaching a plateau in Fig. S1b. On the contrary, within the non-validated subset (322/998, 32%), 134 SNVs exhibited MAFs in NAVARREsel that did not exceed a 2-fold difference in NAVARREval, indicating close MAF between both datasets (Fig. S2). This exploration of MAF patterns ensures a comprehensive understanding of the genetic landscape within the Navarre population and its stability across different datasets.

### Characterization of common Navarrese variants

The annotation of the 676 common Navarrese SNVs revealed 227 synonymous, 371 missense, and five loss-of-function (LoF) variants. These LoF variants were not reported in ClinVar database^12^ and were located in five distinct genes without an associated phenotype, according to OMIM.^13^ Following the ACMG guidelines for variant classification, four were classified as variants of uncertain significance (VUS) and one as benign.^15^

Clinically, 264/676 reported in ClinVar: 1/264 as a risk factor, 181/264 as benign/likely-benign, 32/264 as VUS, 48/264 as having conflicting interpretations, and 2/264 as likely-pathogenic. These likely-pathogenic missense variants were in *SCNN1B* [c.1688G>A:p.Arg563Gln; MAF_NAVARREsel_=0.013, MAF_NAVARREval_=0.016] and in *PTGIS* [c.824G>A:p.Arg275Gln; MAF_NAVARREsel_=0.013, MAF_NAVARREval_=0.021], associated with “low renin hypertension” and “childhood-onset schizophrenia” respectively, according to ClinVar.^12^ The evidence supporting these associations is limited, with a score of 1 out of 4 and reviewed by a single submitter record. Additionally, given its notable prevalence in the Navarrese population, observed in healthy and affected (not related to this phenotype) individuals, these variants might be reconsidered and reclassified as VUS under the ACMG guidelines.^15^

Moreover, common Navarrese variants showed no impact on drug metabolism/efficacy, according to PharmGKB.^14^ Further in silico and enrichment analysis are detailed in Supplementary information.

### Refining disease-causing variant identification in the Navarrese population

We identified common variants in the Navarrese population, highlighting population-specific importance in advancing personalized medicine. The aim was to improve the identification of disease-causing variants during genetic diagnosis using NGS. Therefore, we selected 127 WGS Navarrese patients from *NAGEN1000* project diagnosed with rare disorders and extracted exonic SNVs on chromosomes 1 to 22, averaging 8,871 variants per patient.

We refined the variant list by excluding common variants from *spain*, *1KGP_noIBS*, gnomAD, and Navarre. The Navarrese filtering emerged as the most stringent, resulting in 2.1% of the initial set, compared to 2.7% with gnomAD, 2.9% with *spain* frequencies, and the least restrictive, 4.9% with *1KGP_noIBS* (Fig. 3). This underscores the effectiveness of Navarrese-specific filter in prioritizing and streamlining genetic investigations.

## DISCUSSION

In this study, we aimed to enhance diagnostic precision in the Navarrese population by exploring common population-specific variants. Utilizing WGS data from 358 individuals of Navarre, we identified 61,410 SNVs, with 21,174 common. Genetic analysis shows affinity with European populations and low differentiation with Spanish populations.

Focusing on exclusively common variants in Navarre compared with referenced populations, we obtained 1,069 SNVs, of which 676 were validated in another Navarrese cohort. Of these, none showed clinical or pharmacological relevance beyond what was observed in the Spanish population.^19^ This aligns with the expectation that common population variants are less likely to be associated with disease ethology.

Our findings underscore the relevance of considering population-specific factors in genomic diagnostics, which provides complementary insights alongside pangenome references.^20^ In conclusion, by identifying and excluding common variants within the Navarrese population, we have successfully refined the identification of potential disease-causing variants, contributing to the advancement of personalized medicine for individuals from Navarre. Further research will enhance these insights for broader applications.

## Data Availability

The data that support the findings of this study are available from the corresponding author upon reasonable request.

## DATA AVAILABILITY

Data is available from the corresponding author upon reasonable request.

## FUNDING

*NAGEN1000* and *PharmaNAGEN* were supported by Navarra Gov (Dirección General de Industria, Energia y Proyectos Estrategicos S3). GRANTS_NUMBERS: 0011-1411-2017-000032, 0011-1411-2018-000047.

## AUTHOR INFORMATION

### Contributions

Conceptualization: AM, AA, DGC; Clinical and sample collection: MM, LDM, OT, JS, MAR, SPS; Formal analysis: AM, RC, JPF, VA, DLL, MPC; Data curation: AM; Investigation: AM, EH, MAR, EU, DGC; Funding acquisition: JB, AA; Visualization: AM, EH, MAR, EU; Writing-original draft: AM, EH, MAR, EU, DGC; Writing-review & editing: AM, MAR, EH, EU, SB, SPS, IG, JD, IL, JB, AA, DGC.

### Corresponding authors

Correspondence to Ángel Alonso & David Gomez-Cabrero

### NAGEN scheme

| Name | Department | NAGEN project |
| --- | --- | --- |
| Anda Apiñaniz,<br>Emma | Servicio de Endocrinología y<br>Nutrición, HUN | NAGEN 1,000 |
| Artigas López,<br>Mercedes | Servicio de Genética Médica,<br>HUN | NAGEN 1,000 |
| Bandrés Elizalde, Eva | Servicio de Hematología, HUN | NAGEN 1,000 |
| Basurte Elorz, M<sup>a</sup> Teresa | Servicio de Cardiología, CHN | NAGEN 1,000 |
| Brennan, Paul | NENC NHS Genomic Medicine Centre. Newcastle upon Tyne, UK. | NAGEN 1,000 |
| Celaya Lecea, Concepción | Subdirección de Farmacia, SNS-O | PharmaNAGEN |
| Cuesta Zorita, Manuel Jesús | Servicio de Psiquiatría, Salud mental | NAGEN 1,000 |
| Curi Chercoles, Sergio Miguel | Servicio de Neumología, HUN | NAGEN 1,000 |
| De la Cruz Sánchez, Susana | Servicio de Oncología Médica, HUN | NAGEN 1,000 |
| Erviti López, Juan | Subdirección de Farmacia, SNS-O | PharmaNAGEN |
| Fanlo Mateo, Patricia | Servicio de Medicina Interna, HUN | NAGEN 1,000 |
| González, Luis Angel | AVANTIA 400+. | NAGEN 1,000 |
| Gonzalo Etayo | Navarra de Servicios y Tecnología NASERTIC. Spain | NAGEN 1,000 |
| Gorricho Mendivil, Javier | Subdirección de Farmacia, SNS-O | PharmaNAGEN |
| Guerra Lacunza, Ana | Servicio de Aparato Digestivo, HUN | NAGEN 1,000 |
| Gut, Ivo | Centro Nacional de Análisis Genómicos CNAG. Spain | NAGEN 1,000 |
| Ibáñez Bosch, Rosario | Servicio de Endocrinología y Nutrición, HUN | NAGEN 1,000 |
| Jiménez, Jorge | Navarra de Servicios y Tecnología NASERTIC. Spain | NAGEN 1,000 |
| Lasheras, Gorka | Navarra de Servicios y Tecnología NASERTIC. Spain | NAGEN 1,000 |
| Lorea Bueno | Pharmamodelling | PharmaNAGEN |
| Maite Sarobe Carricas | Servicio de Farmacia Hospitalaria, HUN | PharmaNAGEN |
| Mendioroz Iriarte, Maite | Servicio de Neurología, HUN | NAGEN 1,000 |
| Molinuevo Ruiz de Zarate, José Ignacio | Servicio de Oftalmología, HUN | NAGEN 1,000 |
| Montes Díaz, Marta | Servicio de Anatomía Patológica, HUN | NAGEN 1,000 |
| Navarro, Adela | Servicio de Cardiología, HUN | PharmaNAGEN |
| Onintza Sayar | Pharmamodelling | PharmaNAGEN |
| Pinillos, Iñaki | Navarra de Servicios y Tecnología NASERTIC. Spain | NAGEN 1,000 |
| Purroy Irurzon,<br>Carolina Eugenia | Servicio de Nefrología, HUN | NAGEN 1,000 |
| Sagaseta de Ilurdoz<br>Uranga, M<sup>a</sup> Josefa | Servicio de Pediatría, HUN | NAGEN 1,000 |
| Santesteban<br>Muruzabal, Raquel | Servicio de Dermatología,<br>HUN | NAGEN 1,000 |
| Vicuña, Miren | Servicio de Digestivo, HUN | PharmaNAGEN |
| Viguria, M<sup>a</sup> Cruz | Servicio de Hematología,<br>HUN | PharmaNAGEN |
| Yoldi Petri, M<sup>a</sup><br>Eugenia | Servicio de Pediatría, HUN | NAGEN 1,000 |
| Zubicaray Ugarteche,<br>José Jacinto | Servicio de<br>Otorrinolaringología, HUN | NAGEN 1,000 |
| Zudaire, Maite | Servicio de Hematología,<br>HUN | PharmaNAGEN |

## ETHICS DECLARATIONS

### Competing interests

The authors declare no competing interests.

### Ethics approval

*NAGEN1000* and *PharmaNAGEN* were approved by the Navarra Ethics Committee for Clinical Research (CEIC Navarra).

---

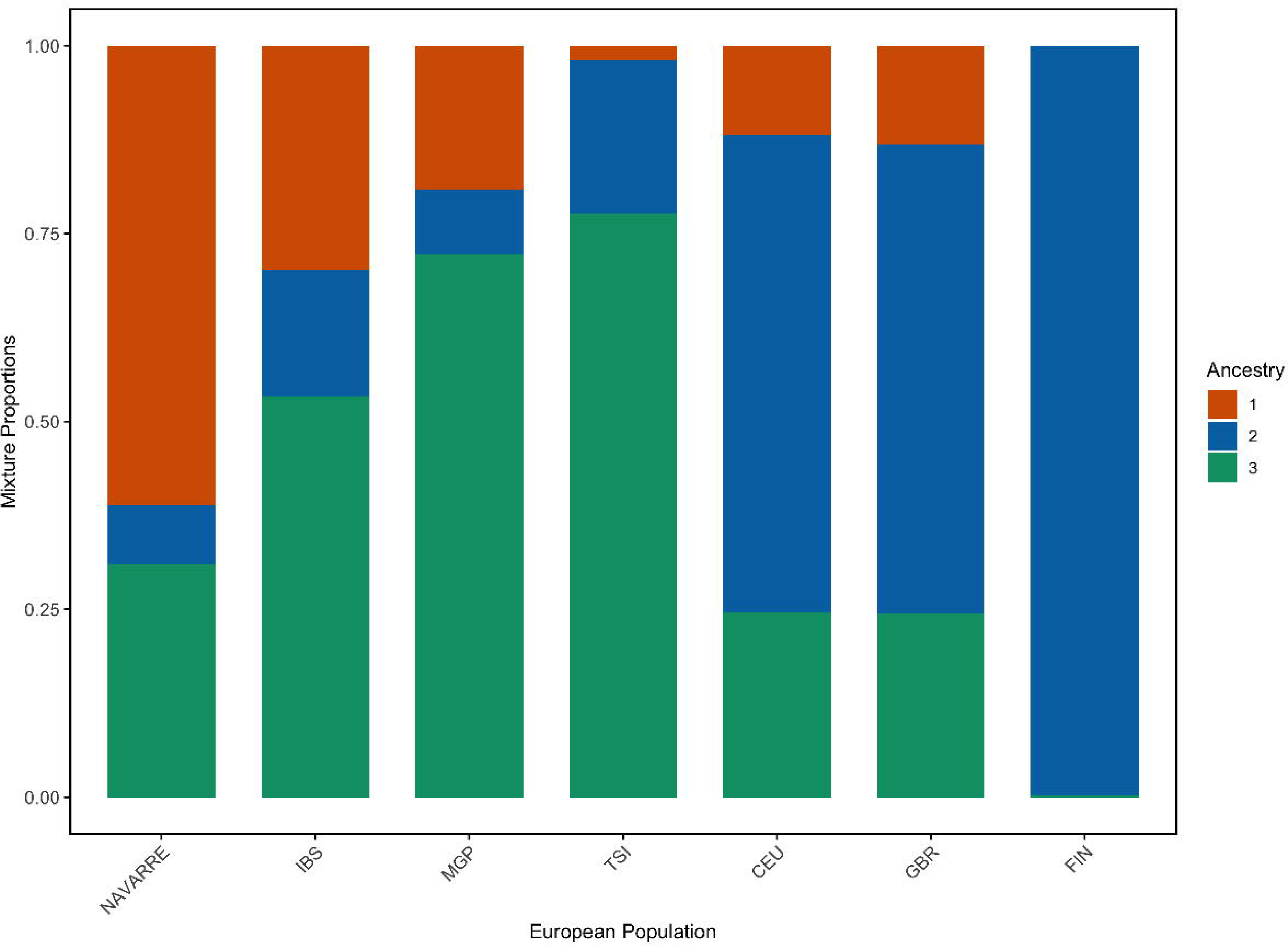

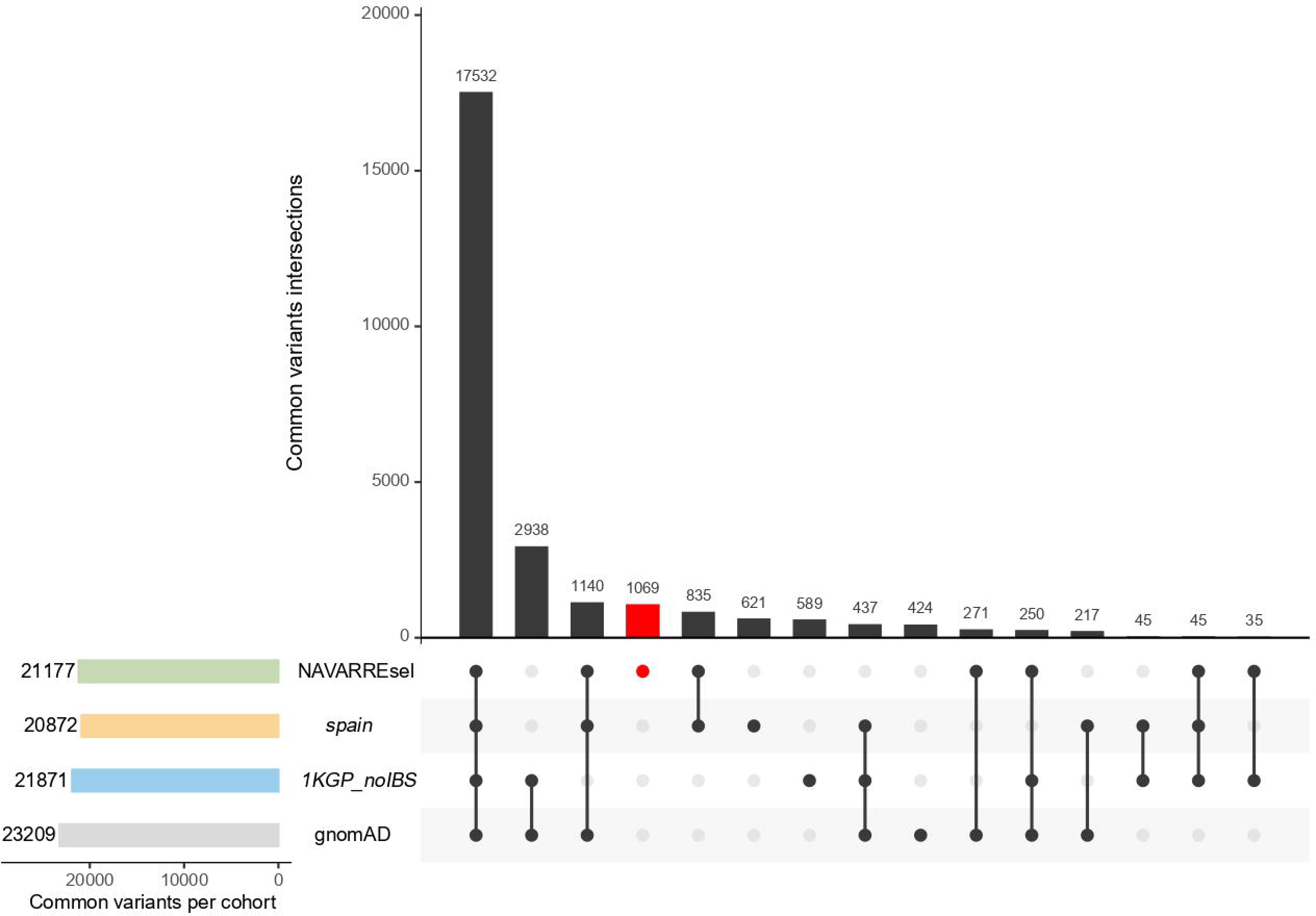

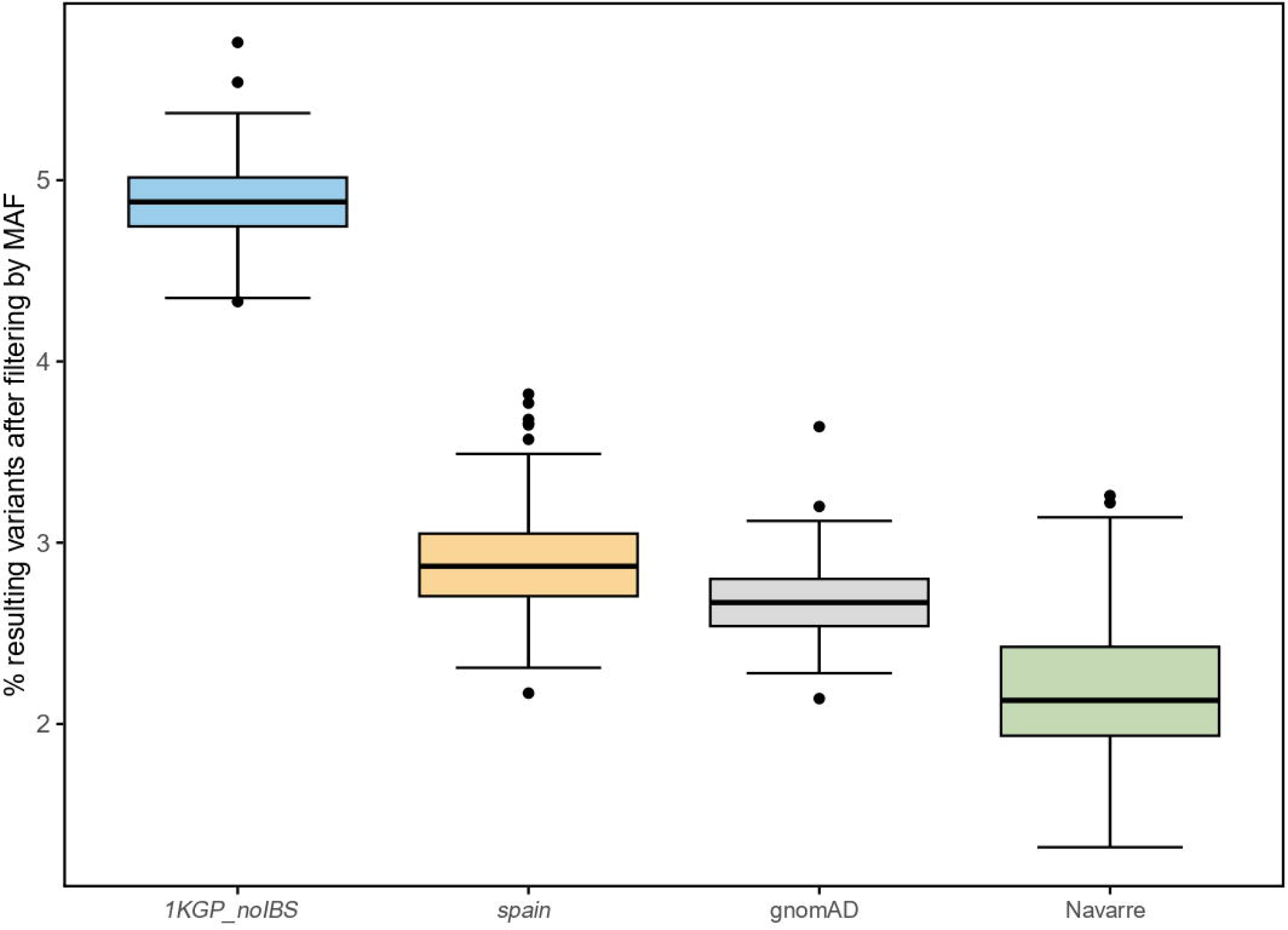

## Notes

### Competing Interest Statement

The authors have declared no competing interest.

### Funding Statement

NAGEN1,000 and PharmaNAGEN were supported by Navarra Gov (Direccion General de Industria, Energia y Proyectos Estrategicos S3). GRANTS_NUMBERS: 0011-1411-2017-000032, 0011-1411-2018-000047.

### Author Declarations

NAGEN1,000 and PharmaNAGEN were approved by Navarra Ethics Committee for Clinical Research (CEIC Navarra).

### Summary of Updates

Reducing the number of words and adding admixture results.

