## Supplementary Information for "Characterization of the common genetic variation in the Spanish population of Navarre"

#### Supplementary Figures

**Figure S1:** Accumulative number of new variants contributed by individuals **a)** All common variants in NAVARREsel (21174 SNVs). **b)** The 676 variants exclusively common in NAVARREsel and validated in NAVARREval.

**Figure S2:** Comparison of MAF between NAVARREsel and NAVARREval encompassing both validated and non-validated variants, with a total of 998 SNVs.

#### Supplementary Tables

**Table S1:** Table of $F_{st}$ values between Navrre against MGP and 1KGP populations.

**Table S2**: Variants' information of the 676 exclusively common SNVs of the Navarre population.

**Figure S1**

**a**

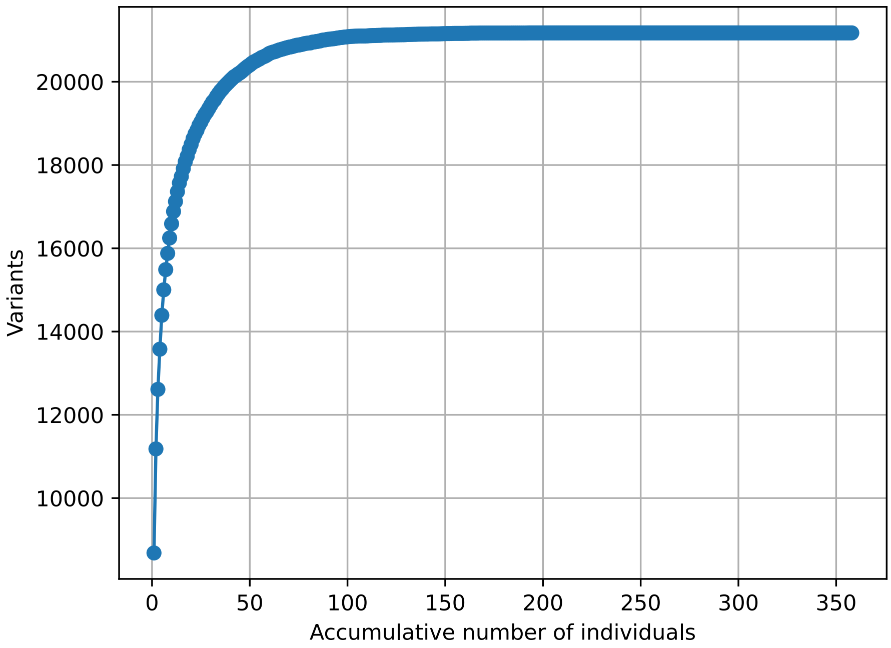
20000
18000
16000
14000
12000
10000
Variants
0
50
100
150
200
250
300
350
Accumulative number of individuals

**b**

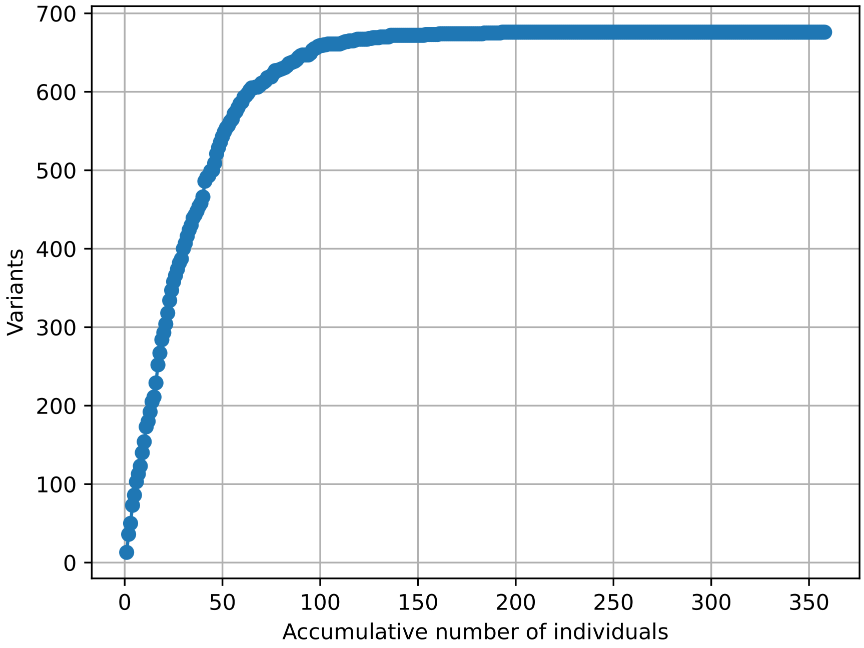
700
600
500
400
300
200
100
0
Variants
0
50
100
150
200
250
300
350
Accumulative number of individuals

**Figure S2**

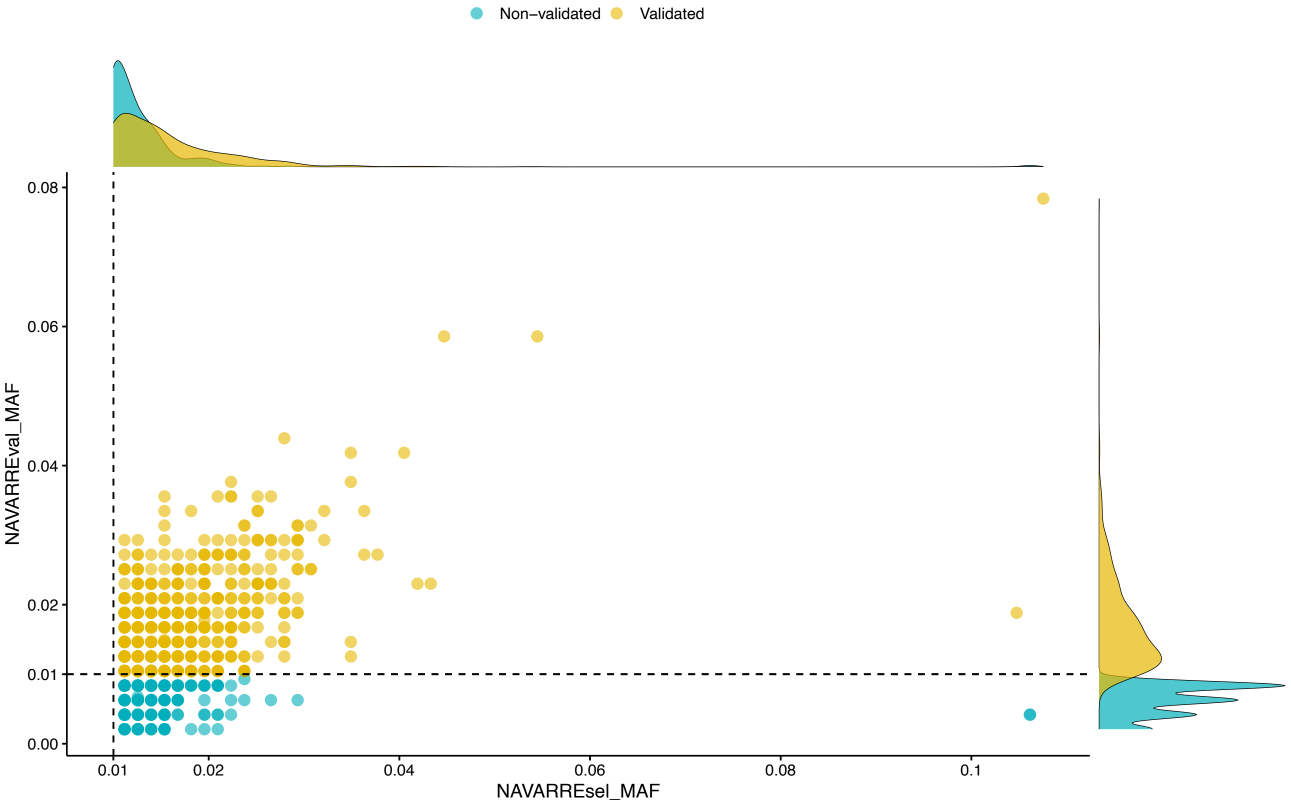

Non−validated
Validated
0.08
0.06
0.04
0.02
0.01
0.00
NAVARREval_MAF
0.01
0.02
0.04
0.06
0.08
0.1
NAVARREsel_MAF

#### MATERIAL AND METHODS

##### *NAGEN1000* and *pharmaNAGEN*

In Navarre, the local Government supports the "*NAGEN scheme*", integrating genomic data analysis into regional healthcare across seven projects. In the *NAGEN1000* project, 688 participants were recruited to uncover the underlying genetic causes of disease using WGS data. These individuals belonged to 294 families, predominantly trios, affected by rare disorders. Likewise, in the *pharmaNAGEN* project, 274 patients with Crohn's disease or ulcerative colitis were recruited to analyze variants related to drug efficacy and toxicity on WES data.[1]

##### Whole genome sequencing and data analysis

High-quality DNA samples from peripheral blood were used in the *NAGEN1000* project to construct short-insert paired-end libraries with an average insert size of 400 bp. DNA fragmentation was performed with Covaris S2 and capillary electrophoresis with Bioanalyzer 2100 (Agilent). Libraries were sequenced on a NovaSeq 6000 (Illumina) with a read length of 2x150 bp. A 30X coverage per sample was targeted. After quality control assessment using FASTQC (https://www.bioinformatics.babraham.ac.uk/projects/fastqc/), the sequenced data was aligned to hs37d5 version of human genome reference GRCh37/hg19 using GEM3.[2] Optical and duplicated reads were flagged with Picard MarkDuplicates (https://broadinstitute.github.io/picard/). Following the established Genome Analysis Toolkit (GATK) best practices pipeline (v3.8),[3] indel realignments and recalibration were applied to the previous BAM files. Variant calling on each sample's BAM file was performed using HaplotypeCaller, with default parameters, resulting in a gVCF file. WGS was conducted at Centro

Nacional de Análisis Genómico (CNAG, Barcelona, Spain) and stored at Navarra de Servicios y Tecnología (NASERTIC, Navarre, Spain).

**Whole exome sequencing and data analysis**

In the *pharmaNAGEN* project, germline DNA was extracted from saliva or blood samples using DNA Blood Maxi Kit (Qiagen) and sequenced with Nextera DNA Exome kit. The raw data was aligned to GRCh37/hg19 genome, sourced from UCSC (https://genome.ucsc.edu/), utilizing BWA.[4] The resulting BAM files were marked using Picard (https://broadinstitute.github.io/picard/). Utilizing GATK v4.1.0,[3] an updated version, eliminates the need for the indel realignment step. Recalibration and variant calling were executed with BQSR and Haplotype tools. This process yielded the final gVCF file for each sample. WES was conducted at CNAG and stored at NASERTIC.

**Individual selection**

Selection criteria included unrelated individuals and Spanish ancestry. To assess relatedness among individuals, identity-by-descent (IBD) was calculated using the method of moments (MoM) with the R package SNPRelate.[5] For validation of ethnicity, the CSVS tool[6] was used to determine the degree of alignment of a sample with the genetic variability of the Spanish population. Individuals with a score equal to or higher than 0.9 were categorized as being of Spanish ancestry.

Applying these selection criteria to the *NAGEN1000* project resulted in a cohort of 358 participants denominated NAVARREsel. Within this group, 127 individuals had been diagnosed with various monogenic diseases, and the most prevalent conditions were polycystic kidney disease (14/127), breast cancer (10/127), and hereditary ataxia (8/127).

Regarding the validation dataset, named NAVARREval, the same inclusion criteria were applied to the *pharmaNAGEN* project, resulting in 239 participants with Crohn's disease (153/239) and/or ulcerative colitis (86/239).

**Variant quality control and filtering in NAVARREsel**

A 358 multi-sample gVCF was generated with the NAVARREsel cohort, and biallelic SNVs located in exonic regions were selected. The targeted exonic interval was extracted from Nextera (https://support.illumina.com/sequencing/sequencing_kits/nextera-dna-exome/downloads.html, downloaded on September 27, 2023). Variants with a read depth of less than 10, a genotype-quality score below 50, or a call rate of less than 100% were removed. Variants on the X and Y chromosomes were eliminated to avoid sex bias, as well as the mitochondrial chromosome, given its complexity. Hardy-Weinberg equilibrium (HWE) score[7] was calculated by PLINK (-hardy), and SNVs significantly deviated with p-value $< 10^{-5}$ were excluded.

**Variant quality control and filtering in NAVARREval**

Genotype information for specific SNVs from NAVARREval samples was extracted using the SelectVariants tool of GATK v4.1.0.[3] SNVs with a call rate lower than 80% and those exhibiting a significant deviation from HWE (p-value $< 10^{-5}$) were excluded from validation.

#### Variant annotation

Variants were annotated using ANNOVAR (version available on October 24, 2019, https://annovar.openbioinformatics.org/).[8] The identification of known variants was performed using the dbSNP database (version GCF_000001405.25, downloaded from https://ftp.ncbi.nih.gov/snp/latest_release/VCF/).[9] SNVs were also annotated: 1) for clinical significance by referring to ClinVar (v.20230930, https://www.ncbi.nlm.nih.gov/clinvar/),[10] OMIM (downloaded on October 2, 2023, https://www.omim.org/),[11] VarSome,[12] and Franklin;[13] and 2) for pharmacological relevance using PharmGKB (https://www.pharmgkb.org/).[14] Additionally, the pathogenicity of the variants was evaluated using CADD (v1.6, https://cadd.gs.washington.edu/),[15] REVEL score (v1.3, https://zenodo.org/records/7072866),[16] spliceAI,[17] and Polyphen2 (http://genetics.bwh.harvard.edu/pph2/dokuwiki/downloads downloaded on October 4, 2023).[18] Variants associated with Crohn's disease and/or ulcerative colitis were identified by referencing the Inflammatory Bowel Disease (IBD) database (accessed on October 10, 2023, from https://www.cbrc.kaust.edu.sa/ibd/index.php?p=ibd#).[19]

#### Population projects

The MGP project was a Spanish initiative, primarily featuring WES data from 267 healthy and unrelated participants, mainly from Andalusia and Galicia (Spanish regions).[20]

The population data from 1KGP phase 3 encompassed 2,504 genome samples representing 26 populations.[21] European populations within this dataset included 503 samples from Europe (EUR), such as British in England and Scotland (GBR), Finnish in Finland (FIN), Iberian population in Spain (IBS), Toscani in Italy (TSI), and Utah residents with Northern and Western European ancestry (CEU). Additionally, there were 504 samples from East Asia (EAS), 489 from South Asia (SAS), 661 from Africa (AFR), and 347 from America (AMR), covering various populations.

The gnomAD genomes project v2.1.1 comprises 15,691 genomes and represents diverse populations worldwide, including Africans, Americans, Asians, and Europeans. Within European populations, the majority originated from North-western, Estonian, Finnish, and a smaller representation from Southern Europe.[22]

**Population frequencies**

Based on the population projects described in the previous section, three populations were generated for this study as reference: 1) gnomAD, with the original frequency from the gnomAD genome project; 2) *1KGP_noIBS*, with the mean of all 1KGP population's frequencies, excluding the IBS cohort of 107 individuals; 3) *spain*, combining the frequencies of the IBS and MGP cohorts. The integration process for the *spain* population consisted of adding the number of total alternate alleles divided by the sum of the total number of alleles across the two cohorts.

**Principal Components Analysis, Admixture, and $F_{ST}$ analysis**

The original VCF files for each chromosome from the 1KGP phase3 were downloaded on September 27, 2023, from https://ftp.1000genomes.ebi.ac.uk/vol1/ftp/release/20130502/. Subsequently, these files were merged using the Picard MergeVCFs tool. Access to the raw data from MGP was granted upon request, and the multi-sample VCF was retrieved for the EGA repository at https://ega-archive.org/datasets/EGAD00001003101. Then, the VCF files from 1KGP, MGP, and NAVARREsel were combined using the GATK tool's CombineGVCFs function. The resulting combined file was used to perform Principal Component Analysis (PCA) with R libraries SNPRelate and SNPassoc, conduct ADMIXTURE (v1.3.0)[23] from 3 to 7 genetic components (K), and calculate the mean pairwise $F_{ST}$ values between populations using vcftools (v0.1.17).[24]

#### Enrichment analysis

Functional enrichment, including pathway (KEGG), biological process (GO), disease (OMIM), and human phenotype ontology (HPO), was performed using WebGestalt (https://www.webgestalt.org/).[25]

### RESULTS

**In silico predictors and enrichment analysis**

Analysis of the 676 variants with *in silico* functional predictors revealed eight variants as pathogenic by three different pathogenicity tools (REVEL_score > 0.8,[16] CADD > 20,[15] and Polyphen indicating “probably” or “possibly”)[18] However, a comprehensive examination of clinical databases, including ClinVar,[10] Varsome,[12] Franklin,[13] contradicted these predictions based on ACMG criteria. Instead, the majority of these variants were classified as uncertain significance (1/8), likely benign (5/8), and benign (2/8). According to the ACMG classification, these variants were not disease-related.[26]

Additionally, the variant c.387-1G>T in *BPIFB3* gene ($MAF_{NAVARREsel}$=0.0154, $MAF_{NAVARREval}$=0.0209), predicted to impact the canonical splicing acceptor site (spliceAI score = 0.99), was reclassified as benign based on the allele frequency in Navarre population, and the ACMG criteria.[17]

Enrichment analysis revealed no statistical significance (p-value < 0.05) in pathways, biological processes, related diseases, or phenotypic ontologies.[25]
